# Virtual Noncontrast Imaging with Deep-Silicon Photon-Counting CT: Matched Phantom and Initial Human Evaluation

**DOI:** 10.64898/2026.10.02.26364524

**Authors:** Navid Azimi, Arnaud Brian-Choux, Samuel Feemster, Alireza Rahbar, Hugo Linder, Dominic Crotty, Zhye Yin, Patricia Balthazar, Amir Pourmorteza

## Abstract

**Objectives:** To compare virtual noncontrast (VNC) error between deep-silicon photon-counting CT (PCCT) and dual-energy energy-integrating detector CT (EID-CT) in a matched phantom and characterize tissue-specific error in humans.

**Materials and methods:** This prospective exploratory study included a matched phantom experiment and pilot human evaluation. Prototype projection-domain water-density maps served as the VNC surrogate. Twenty-four vials containing 0.1–20 mg I/mL in deionized water or simulated blood were scanned with PCCT and EID-CT. Three men underwent true noncontrast and contrast-enhanced PCCT; 80 matched measurements were analyzed. Paired t tests and mixed-effects models assessed system differences and iodine associations.

**Results:** Three participants (mean age, 67 ± 10 years; all men) were included. Mean VNC error was 29.1 HU lower with PCCT than EID-CT (95% CI, 15.5–42.7 HU; *p* < 0.001). Error was not associated with iodine concentration for PCCT (slope, 0.03 HU per mg I/mL; 95% CI, −0.92 to 0.98; *p* = 0.95) but increased for EID-CT (slope, 3.51 HU per mg I/mL; 95% CI, 2.56–4.46; *p* < 0.001). The slopes differed by 3.48 HU per mg I/mL (*p* < 0.001). In humans, errors in five non-blood-pool tissues were within 5 HU. Error was not associated with iodine concentration among 61 non-blood-pool measurements (adjusted slope, −0.17 HU per mg I/mL; 95% CI, −1.22 to 0.88; *p* = 0.75).

**Conclusion:** PCCT provided VNC measurements with errors closer to zero and less dependent on iodine than EID-CT in the phantom. In humans, error was generally small in non-blood-pool tissues and was not associated with iodine concentration.

**Key Points:** *Question:* Can deep-silicon photon-counting CT provide virtual noncontrast measurements with errors close to zero and little iodine dependence in a matched phantom and human tissues?

*Findings:* Deep-silicon photon-counting CT showed phantom errors closer to zero and less dependent on iodine than dual-energy EID-CT; errors were generally small in non-blood-pool tissues.

*Clinical Relevance Statement:* Deep-silicon photon-counting CT provided virtual noncontrast measurements with little iodine dependence in non-blood-pool tissues, supporting evaluation in larger cohorts as an alternative to separate true noncontrast acquisitions.

## Introduction

Virtual noncontrast (VNC) images may eliminate a separate true noncontrast (TNC) acquisition while remaining inherently aligned with the corresponding contrast-enhanced images, an important advantage in cardiac and abdominal imaging [1].

Current commercial photon-counting CT (PCCT) systems use cadmium-telluride (CdTe) detectors in a compact face-on geometry and acquire spectral data using two to four energy thresholds [2; 3]. The system evaluated here instead uses deep-silicon (dSi) detectors in a thicker edge-on geometry and records eight energy bins [3–7]. These differences in detector material, geometry, and spectral sampling may affect material decomposition; therefore, VNC performance should be evaluated separately for the two detector designs. An initial human study demonstrated the feasibility of VNC with the dSi PCCT system, but its performance across controlled iodine concentrations and different human tissues remains incompletely characterized [8].

The purpose of this study was to compare VNC error between dSi PCCT and dual-energy CT with an energy-integrating detector (EID-CT) across controlled iodine concentrations in a phantom and to characterize VNC error and its association with iodine concentration in humans. We hypothesized that dSi PCCT would provide VNC errors closer to zero and less dependence on iodine concentration than dual-energy EID-CT.

## Materials and Methods

### Study Oversight and Role of Sponsor

This prospective human study was approved by the Swedish Ethical Review Authority and conducted in accordance with the Declaration of Helsinki. All participants provided written informed consent. This work was performed under a sponsored research agreement between GE HealthCare and Emory University. Authors who were neither employees of nor consultants for GE HealthCare had complete control of the data and analyses and the decision to submit the manuscript for publication.

### Experimental Design

This exploratory technical-development study comprised complementary phantom and human evaluations. The matched phantom experiment compared VNC error between deep-silicon PCCT and rapid kilovoltage-switching energy-integrating detector CT across known iodine concentrations in deionized water and simulated blood. The prospective human evaluation characterized tissue-specific VNC error and assessed its association with measured iodine concentration.

### Phantom Construction and Test Object

A 200-mm-diameter modular CT phantom with removable inserts consisting of 2-mL microcentrifuge vials (Thermo Fisher Scientific) in a three-dimensional printed housing filled with synthetic 20% ballistic gel (Clear Ballistics, Greenville, SC) was used. For this study, 24 vials contained iohexol (Omnipaque 350, GE HealthCare, Chicago, IL) diluted in either deionized water or water containing ferrous sulfate (Lannett Company, Trevose, PA) heuristically adjusted to approximately 60 HU at 70 keV to simulate blood. Duplicate iodine-water vials were prepared at 0.1, 0.2, 0.5, 1, 2, 5, 10, 15, and 20 mg I/mL, and duplicate iodine-simulated-blood vials were prepared at 5, 10, and 17.5 mg I/mL. Additional reference vials containing deionized water or simulated blood were included (Figure 1).

**Figure 1.**
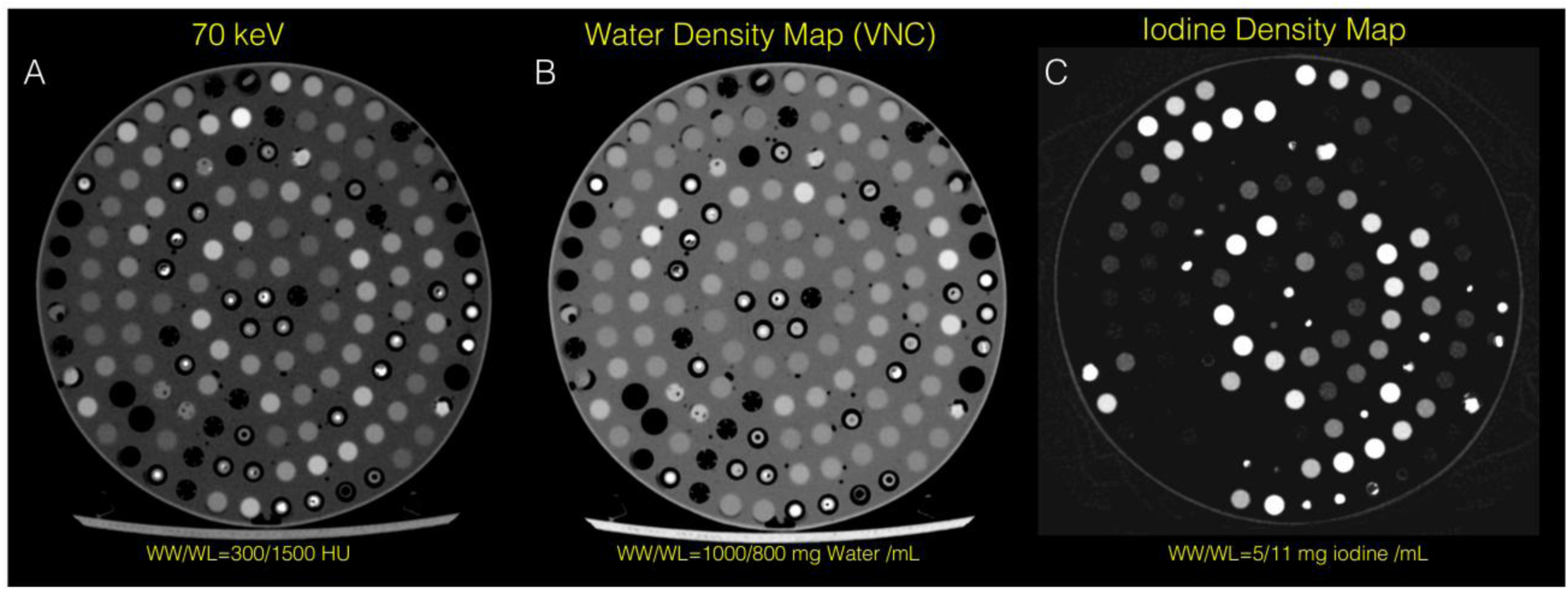
Phantom test object imaged with deep-silicon PCCT. (a) 70-keV virtual monoenergetic image, (b) water-density map, and (c) iodine-concentration map. The test object contained vials with iodine in deionized water or simulated blood within a 20-cm holder surrounded by 20% ballistic gel. The water-density map was used for VNC measurements. PCCT = photon-counting CT; VNC = virtual noncontrast surrogate derived from the water-density map.

### Human Subjects

Three asymptomatic adult volunteers were prospectively and consecutively enrolled in September 2024 in Karolinska University Hospital (Stockholm, Sweden). Exclusion criteria were renal failure or known allergy to iodinated contrast material. Because this was an exploratory pilot evaluation, no formal sample size calculation was performed.

### CT Systems and Acquisition Protocols

Phantom imaging was performed on an edge-on-irradiated deep-silicon PCCT system (Photonova Spectra, GE HealthCare) and a rapid kilovoltage-switching dual-energy EID-CT system (Revolution Apex Elite, GE HealthCare). The systems had matched acquisition geometry and identical x-ray tubes. The PCCT system recorded eight energy bins: three dynamically configured for Compton scatter measurements, two were set at K-edge energies of iodine and gadolinium and the rest were evenly distributed. The EID-CT system used rapid 80/140-kVp switching.

Phantom scans were acquired axially at similar radiation output: PCCT at 120 kVp and 300 mAs (volume CT dose index [CTDIvol], 20.51 mGy) and EID-CT at 80/140 kVp and 370 mAs (CTDIvol, 19.44 mGy).

Participants underwent helical 120-kVp chest-abdomen PCCT before and after administration of 175 mL iodixanol (320 mg I/mL) according to clinical routine settings. Automatic exposure control (Smart mA, GE HealthCare) was used with a pitch of 0.99 and rotation time of 0.5 second. All participants underwent arterial-phase imaging, and one also underwent portal-venous imaging. Tube current and CTDIvol were 120±24 mA and 4.1±0.8 mGy precontrast, 238±50 mA and 8.1±1.7 mGy arterial, and 138 mA and 4.69 mGy portal venous.

### Image Reconstruction and Material Decomposition

Phantom images were reconstructed with filtered back projection and a standard body kernel (Stnd); human images were reconstructed with medium-strength deep-learning reconstruction and the same kernel. Section thickness was 0.625 mm for EID-CT and 0.416 mm for PCCT. All images were reconstructed with a 512 × 512 matrix.

Water-density and iodine-concentration maps were generated from the eight-energy-bin data using vendor-provided prototype projection-domain material-decomposition software. The PCCT scanner was FDA-cleared; only the material-decomposition software was prototype. Because a dedicated commercial VNC reconstruction was unavailable, the postcontrast water-density map was used as a surrogate and is referred to as VNC hereafter.

### Quantitative Image Analysis

#### Phantom ROI Analysis

Matched 30-mm² circular ROIs were placed within each vial on the EID-CT and PCCT VNC images. Phantom VNC error was calculated from the mean ROI measurements as the iodine-containing vial value minus the corresponding deionized-water or simulated-blood reference on the same CT system.

#### Participant ROI Analysis

A fellowship-trained abdominal radiologist (P.B., 5 years of post-training experience) placed elliptical ROIs at 80 anatomically matched locations on true noncontrast (TNC) and contrast-enhanced scans. ROIs were at least 20 mm² and covered as much of a uniform region as possible. Locations included blood in arteries or cardiac chambers, subcutaneous and interstitial fat, myocardium, kidneys, liver, skeletal muscle, pancreas, spleen, and incidentally detected hypoenhancing liver or pancreatic lesions. In participants, VNC error was calculated as the mean postcontrast VNC measurement minus the corresponding mean TNC measurement. Iodine concentration was calculated as the difference between the contrast-enhanced and TNC iodine-map measurements and reported in mg I/mL.

VNC error values closer to zero indicated better agreement with the corresponding reference; positive and negative values indicated overestimation and underestimation, respectively.

### Statistical Analysis

Phantom VNC errors were summarized using means and standard deviations. VNC errors for the 24 matched phantom vials were compared between EID-CT and PCCT using a paired t test. The paired difference was calculated as EID-CT VNC error minus PCCT VNC error. The association between iodine concentration and phantom VNC error was assessed using a linear mixed-effects model that included CT system, iodine concentration, and their interaction. The model adjusted for whether iodine was diluted in deionized water or simulated blood and accounted for paired measurements of each vial on both systems. The system-by-iodine interaction tested whether the association differed between systems.

Participant VNC errors were summarized by tissue using medians and interquartile ranges. The association between iodine concentration and VNC error among the 61 non-blood-pool measurements was assessed using a linear mixed-effects model adjusted for participant and acquisition phase. ROI location was included as a random intercept to account for locations measured in both contrast phases.

Effect estimates were reported with 95% CIs. All tests were two-sided, with *p* < 0.05 considered to indicate statistical significance. Analyses were performed using MATLAB, version R2024b (MathWorks).

## Results

### Participant Characteristics

All three enrolled participants (mean age, 67±10 years; all men; mean body mass index, 25.8±6.7 kg/m²) were included and completed TNC and contrast-enhanced PCCT examinations (Figure 3; Electronic Supplementary Material Figures S1 and S2).

### Phantom VNC Error

At the tested iodine concentrations, mean VNC error ranged from −1.6 to 19.0 HU with PCCT and from 11.4 to 84.7 HU with EID-CT (Figure 2). VNC error showed no association with iodine concentration for PCCT (slope, 0.03 HU per mg I/mL; 95% CI, −0.92 to 0.98; *p* = 0.95) but increased with iodine concentration for EID-CT (slope, 3.51 HU per mg I/mL; 95% CI, 2.56–4.46; *p* < 0.001). The slopes differed by 3.48 HU per mg I/mL (95% CI, 2.19–4.77; *p* < 0.001). Across the 24 matched vials, mean VNC error was 29.1 HU lower with PCCT than with EID-CT (95% CI, 15.5–42.7 HU; *p* < 0.001).

**Figure 2.**
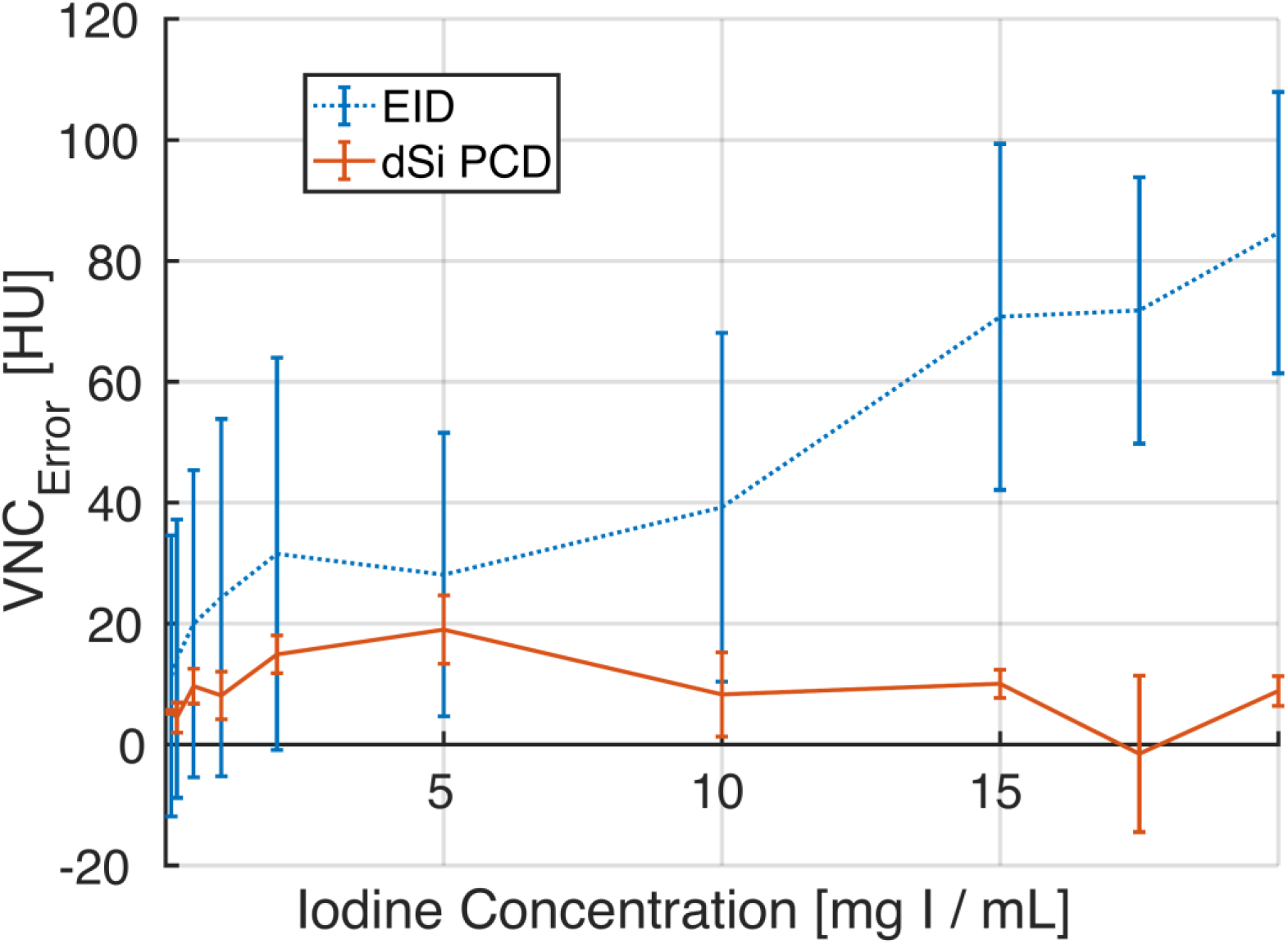
VNC error by iodine concentration for rapid kilovoltage-switching EID-CT and deep-silicon PCCT. VNC error was calculated relative to the corresponding deionized-water or simulated-blood reference on the same CT system. Points represent means, and error bars represent standard deviations. Each concentration included two vials, except 5 and 10 mg I/mL, which each included four. VNC error was not associated with iodine concentration for PCCT (slope, 0.03 HU per mg I/mL) but increased for EID-CT (slope, 3.51 HU per mg I/mL). The slopes differed by 3.48 HU per mg I/mL (95% CI, 2.19–4.77; *p* < 0.001). CI = confidence interval; EID-CT = energy-integrating detector CT; HU = Hounsfield unit; PCCT = photon-counting CT; VNC = virtual noncontrast surrogate derived from the water-density map.

**Figure 3.**
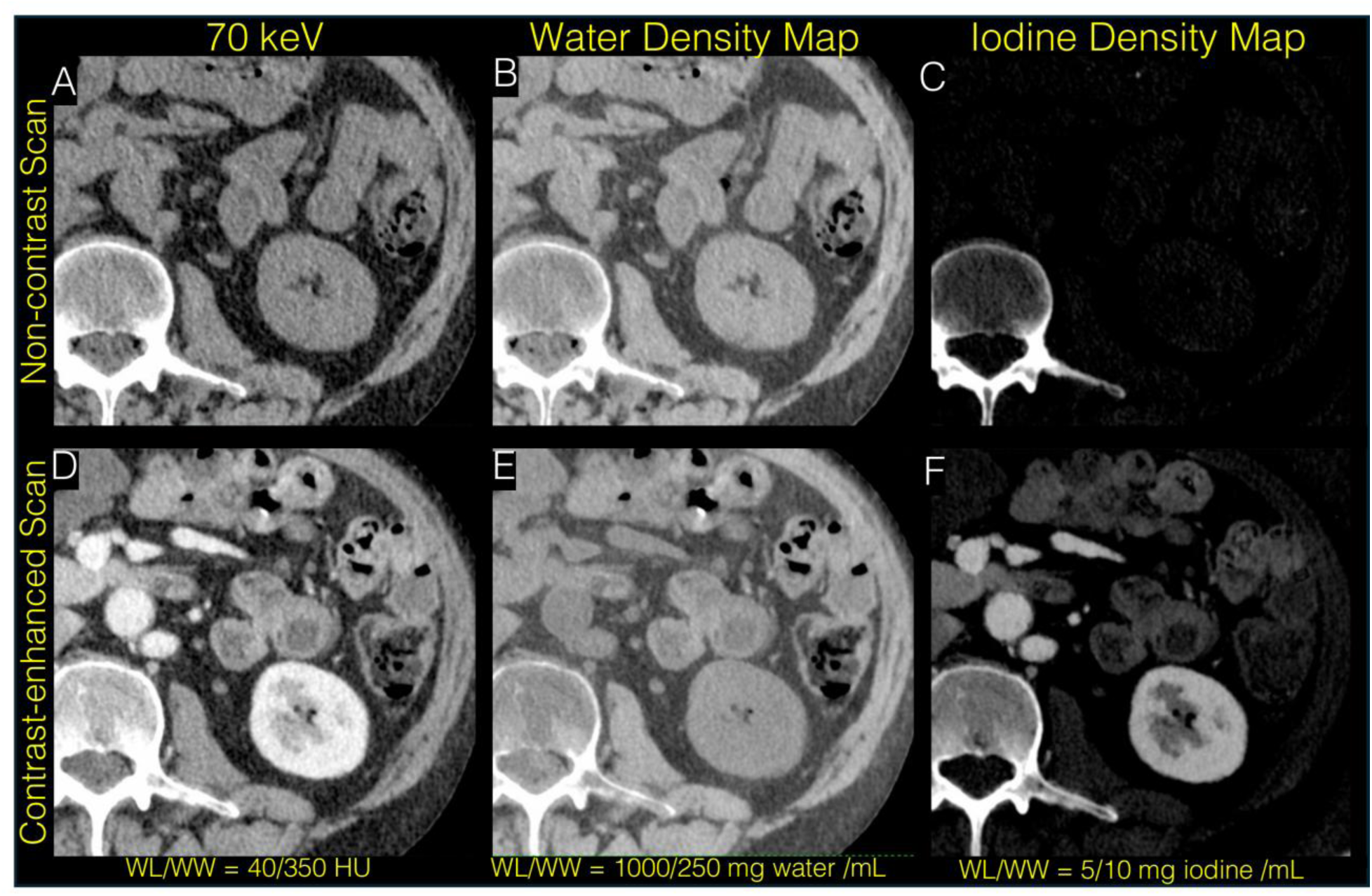
Representative axial deep-silicon PCCT images at the level of the left kidney acquired before (top row) and after (bottom row) administration of iodinated contrast material. (a, d) 70-keV virtual monoenergetic images, (b, e) water-density maps, and (c, f) iodine-concentration maps. The precontrast water-density map (b) was used as the TNC reference, and the postcontrast water-density map (e) was used for VNC measurements. PCCT = photon-counting CT; TNC = true noncontrast; VNC = virtual noncontrast surrogate derived from the postcontrast water-density map.

### Human VNC Error

Eighty matched measurements included 19 blood-pool and 61 non-blood-pool measurements. Median VNC error across all measurements was −6.6 HU (interquartile range [IQR], −12.9 to 0.0 HU). Iodinated blood had the highest iodine concentrations and showed the largest negative median error (−19.6 HU; IQR, −25.6 to −10.8 HU). Among non-blood-pool tissues, liver had the most negative median error (−8.6 HU), followed by kidney (−7.2 HU) and lesions (−6.5 HU). Median VNC error was within 5 HU in pancreas (−0.5 HU), fat (−0.9 HU), myocardium (−0.9 HU), skeletal muscle (−3.0 HU), and spleen (−4.3 HU), including within 1 HU in pancreas, fat, and myocardium (Figure 4a).

**Figure 4.**
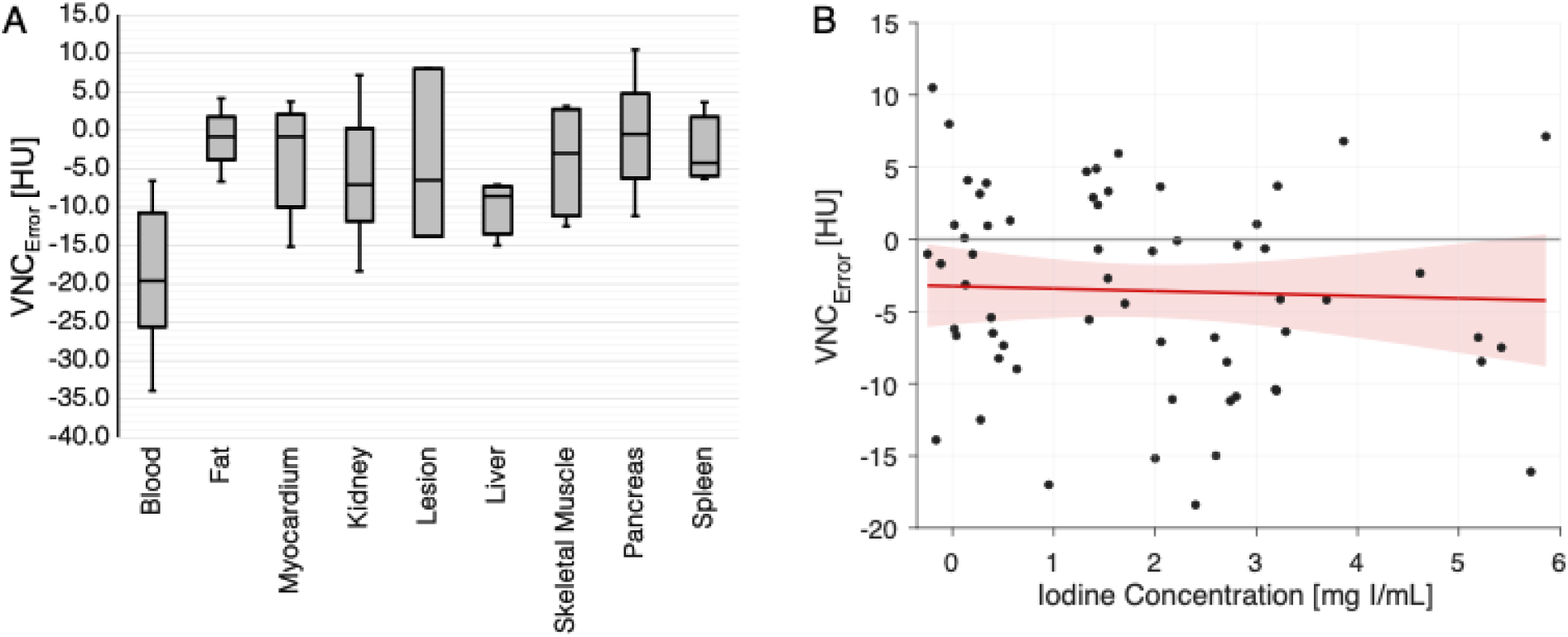
Human VNC error. VNC error was calculated as the postcontrast VNC measurement minus the corresponding TNC measurement. (a) Distribution of VNC error across 80 matched blood-pool and non-blood-pool measurements. Boxes show the interquartile range, center lines show the median, whiskers show the range, and points show individual measurements. (b) Association between iodine concentration and VNC error among 61 non-blood-pool measurements. The solid red line shows the fitted relationship after adjustment for subject and acquisition phase, and the shaded red area shows the 95% CI for the fitted mean. VNC error was not associated with iodine concentration (adjusted slope, −0.17 HU per mg I/mL; 95% CI, −1.22 to 0.88; *p* = 0.75). CI = confidence interval; HU = Hounsfield unit; TNC = true noncontrast; VNC = virtual noncontrast surrogate derived from the water-density map.

### Association between Iodine Concentration and VNC Error

Among the 61 non-blood-pool measurements, VNC error was not associated with iodine concentration (adjusted slope, −0.17 HU per mg I/mL; 95% CI, −1.22 to 0.88; *p* = 0.75) (Figure 4b).

## Discussion

In this exploratory technical-development study, dSi PCCT provided more accurate VNC measurements and showed less dependence on iodine concentration than dual-energy EID-CT in matched phantom vials. In participants, median VNC error was within 5 HU in five tissues and within 1 HU in pancreas, fat, and myocardium. This level of agreement is notable because 5 HU is a commonly used CT quality-control tolerance [9]. VNC error was not associated with iodine concentration but varied among tissues, with more negative error in liver and especially blood.

These findings extend the initial human evaluation of dSi PCCT by Salyapongse et al. [8], which demonstrated the feasibility of VNC in five participants but did not include a controlled phantom or a matched EID-CT comparator. The present study evaluated known iodine concentrations in deionized water and simulated blood, directly compared matched vials on dSi PCCT and dual-energy EID-CT at similar radiation output, and characterized tissue-specific VNC error in participants.

Published studies of current commercial CdTe-based PCCT systems have reported tissue-dependent VNC error, including larger errors in adipose tissue, kidneys, and vascular structures [10–13]. In the present study, VNC error appeared smaller in several non-blood-pool tissues, including fat, myocardium, pancreas, skeletal muscle, and spleen. Comparisons across studies should be interpreted cautiously because acquisition methods, reconstruction and VNC algorithms, participant populations, and ROI methods differed. The eight-bin spectral sampling of dSi could contribute to more accurate material decomposition and less dependence of VNC error on iodine concentration, but these possible explanations were not tested.

Sawall et al. showed that separate TNC and contrast-enhanced acquisitions may be more dose efficient for material decomposition when the two acquisitions can be accurately registered [14]. However, exact registration between separate acquisitions is rarely achievable, particularly in cardiac and abdominal imaging, because of cardiac and respiratory motion and changes in organ position and shape. VNC remains inherently aligned with the corresponding contrast-enhanced images and is therefore clinically important even when separate acquisitions may have a technical advantage under ideal registration.

Iodinated blood, which contained the highest iodine concentrations, showed the largest negative VNC error and illustrates a fundamental limitation of iodine-based VNC. Iodine and soft tissue have similar energy-dependent x-ray attenuation within the diagnostic CT range, which limits their separation and prevents complete elimination of VNC error [1]. Contrast agents with a prominent K edge within this energy range, such as gadolinium, tantalum, or gold, provide a more distinctive spectral signature and may be better suited to material decomposition and VNC imaging [1; 6; 15]. The eight energy bins available with dSi provide greater flexibility for sampling around a K edge than the two to four thresholds available with current commercial CdTe-based systems and may facilitate K-edge imaging [2; 6]. However, K-edge contrast agents are not yet in widespread clinical use for CT.

This study had limitations. The pilot human evaluation included three asymptomatic men from a single institution, limiting the generalizability of the tissue-specific findings. Most contrast-enhanced acquisitions were obtained in the arterial phase, limiting evaluation across contrast phases. However, prior work found that venous-phase VNC agreed more closely with TNC than arterial-phase VNC, particularly in the aorta, making arterial imaging a demanding test [16]. The evaluated water-density maps were generated with prototype material-decomposition software and used as a surrogate for a commercial VNC product; finalized VNC software may include additional calibration, corrections, or postprocessing and may perform differently. The phantom did not reproduce the full composition or motion of human tissues, and the study did not include a reader assessment or clinical diagnostic task.

In conclusion, dSi PCCT provided more accurate VNC measurements than dual-energy EID-CT across the tested iodine concentrations in the phantom. In the pilot human evaluation, VNC error showed little dependence on iodine concentration and was generally small in non-blood-pool tissues, with errors for most tissues meeting the tolerance commonly used for CT number calibration. These promising findings support the clinical utility of dSi PCCT VNC, but further evaluation in larger cohorts is warranted.

## Data Availability

The study data are not publicly available because the human component includes participant data. Requests for access to data and materials may be directed to the corresponding author and will be reviewed individually.

## Abbreviations

CI: confidence interval;
CTDIvol: volume CT dose index;
dSi: deep-silicon;
EID-CT: energy-integrating detector CT;
HU: Hounsfield unit;
IQR: interquartile range;
PCCT: photon-counting CT;
ROI: region of interest;
TNC: true noncontrast;
VNC: virtual noncontrast.

## Declarations

### Ethics approval and consent to participate

The prospective human study was approved by the Swedish Ethical Review Authority and conducted in accordance with the Declaration of Helsinki. Written informed consent was obtained from all participants.

### Consent for publication

Written informed consent for publication of de-identified images and data was obtained from all participants as part of study consent.

### Authors’ contributions

AB-C and NA: data curation, formal analysis, methodology, visualization, and writing – review and editing. SF and AR: data curation, formal analysis, and writing – review and editing. HL and DC: data curation, resources, and writing – review and editing. ZY: data curation, project administration, resources, software, and writing – review and editing. PB: investigation, validation, and writing – review and editing. AP: conceptualization, formal analysis, funding acquisition, investigation, methodology, project administration, resources, software, supervision, validation, writing – original draft, and writing – review and editing.

All authors reviewed and approved the final manuscript.

## Acknowledgements

Not applicable.

