## Supplementary Figures for "Virtual Noncontrast Imaging with Deep-Silicon Photon-Counting CT: Matched Phantom and Initial Human Evaluation"

ELECTRONIC SUPPLEMENTARY MATERIAL

Figure S1.

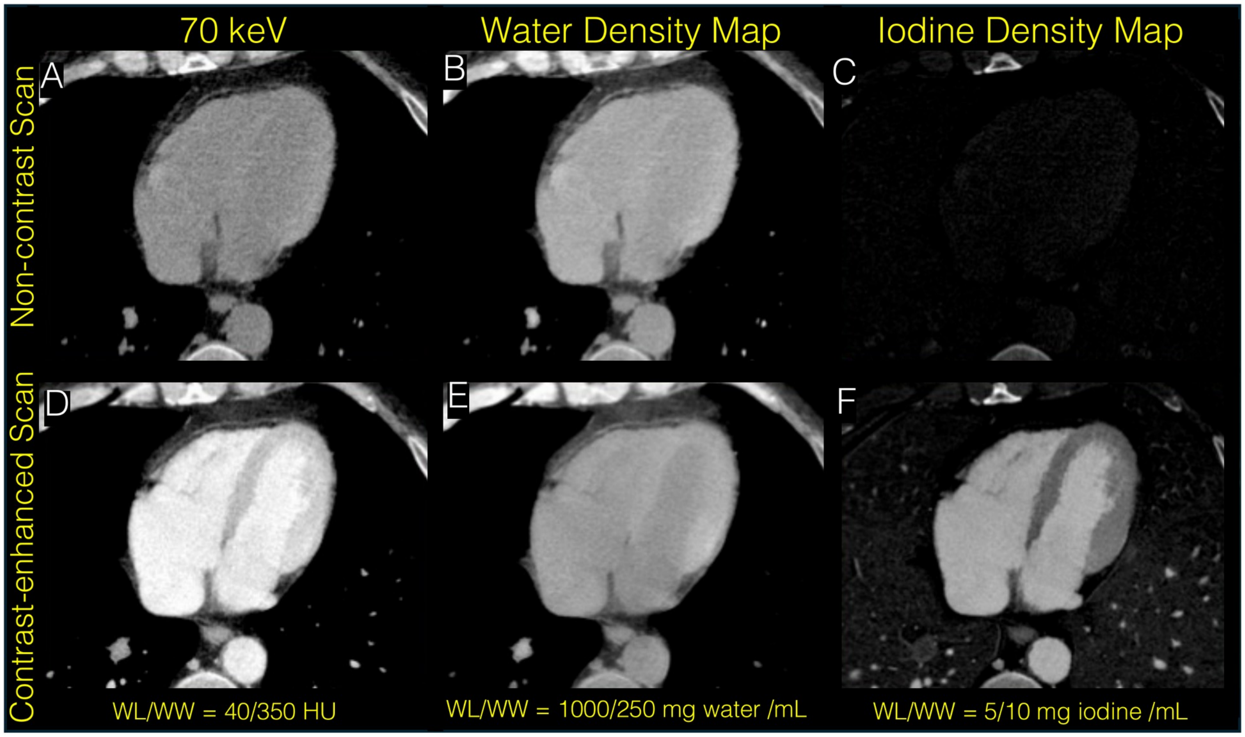


**Figure S1.** Representative axial deep-silicon PCCT images at the level of the heart acquired before (top row) and after (bottom row) administration of iodinated contrast material. (a, d) 70-keV virtual monoenergetic images, (b, e) water-density maps, and (c, f) iodine-concentration maps. The precontrast water-density map (b) was used as the TNC reference, and the postcontrast water-density map (e) was used for VNC measurements. PCCT = photon-counting CT; TNC = true noncontrast; VNC = virtual noncontrast surrogate derived from the postcontrast water-density map.

Figure S2.

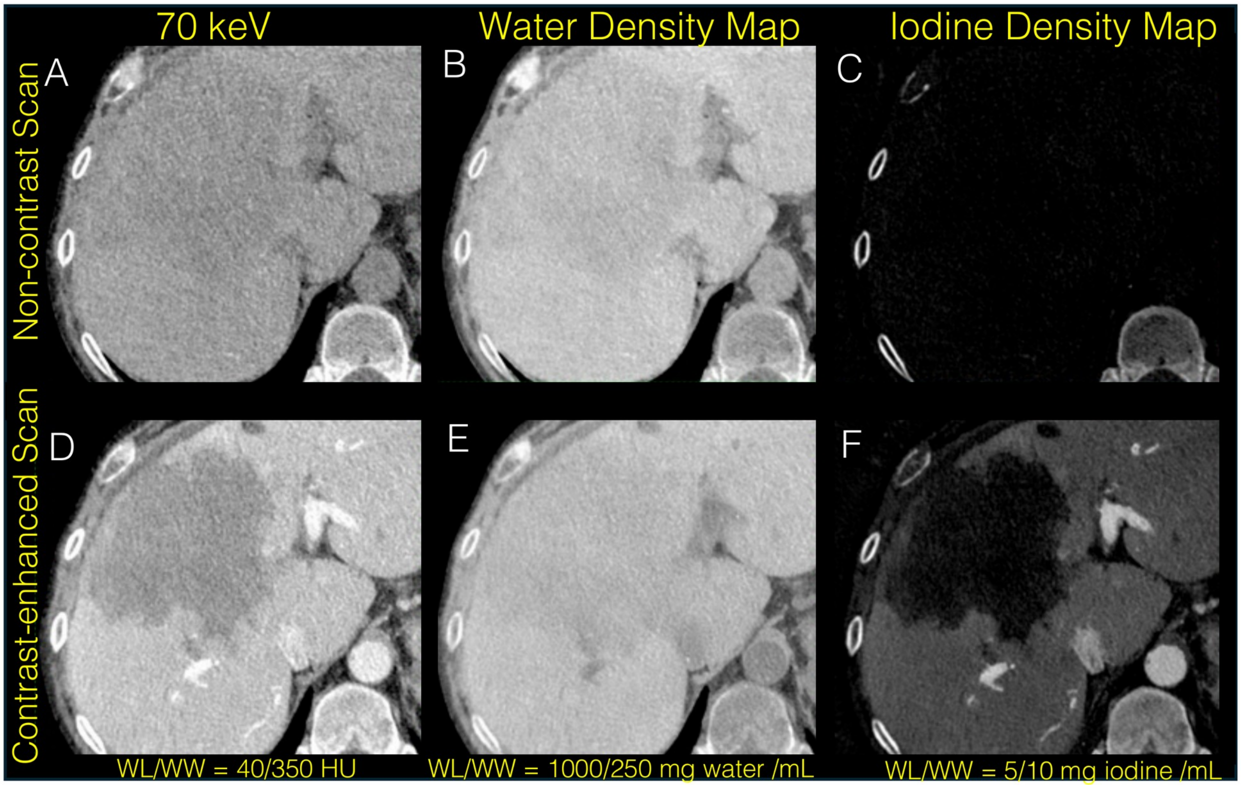


**Figure S2.** Representative axial deep-silicon PCCT images through an incidentally detected hypoenhancing liver lesion acquired before (top row) and after (bottom row) administration of iodinated contrast material. (a, d) 70-keV virtual monoenergetic images, (b, e) water-density maps, and (c, f) iodine-concentration maps. The precontrast water-density map (b) was used as the TNC reference, and the postcontrast water-density map (e) was used for VNC measurements. PCCT = photon-counting CT; TNC = true noncontrast; VNC = virtual noncontrast surrogate derived from the postcontrast water-density map.
